# Genetic liability to hip osteoarthritis confers neurovascular protection against Alzheimer’s disease despite depression-mediated phenotypic comorbidity

**DOI:** 10.64898/2026.03.04.26347509

**Authors:** Qi Xu, Pianpian Zhao, Jianguo Tao, Houfeng Zheng

**Affiliations:** Suzhou Laboratory of Precision Health and Data Science, the Second Affiliated Hospital of Soochow University, Suzhou, Jiangsu 215004, China; The Affiliated Hangzhou First People’s Hospital, School of Medicine, Westlake University, Hangzhou, Zhejiang 310000, China; Center for Biobased Materials, Muyuan Laboratory, Zhengzhou, Henan Province 450016, China

## Abstract

**Background:** The relationship between hip osteoarthritis (hip OA) and Alzheimer’s disease (AD) presents a critical paradox within the emerging "bone-brain axis": widespread phenotypic comorbidity sharply contradicts evolutionary theories of biological antagonism. This study integrates longitudinal and multi-omic analyses to determine whether this clinical overlap masks an underlying genetic neuroprotection.

**Methods:** We analyzed longitudinal phenotypic data from 261,767 UK Biobank participants using Cox proportional hazards and Fine-Gray competing risk models. To investigate the shared genetic architecture, we applied MiXeR modeling to genome-wide association study summary statistics. Causal relationships were evaluated using global and cell-type-stratified Mendelian randomization across eight distinct brain cell types. Shared genomic loci were identified via conjunctional/conditional false discovery rate and fine-mapping. Single-nucleus RNA-sequencing (snRNA-seq) data from the ROSMAP cohort validated the disease-associated transcriptional dynamics of prioritized target genes.

**Results:** Observational survival analyses initially suggested an increased AD risk in patients with hip OA; however, this association was fully attenuated after adjusting for a history of depression, revealing a "phenotypic illusion" driven by the pain-depression axis. Conversely, cell-type-stratified genomic analyses uncovered a profound biological antagonism: genetic liability for hip OA confers robust neuroprotection specifically localized to the neurovascular unit (NVU), primarily driven by astrocytes and pericytes. Mechanistically, this NVU fortification is orchestrated by the *MAPT* locus and PI3K/AKT signaling, with snRNA-seq confirming the active transcriptional remodeling of these core effectors in the AD brain.

**Conclusion:** We demonstrate that genetic liability to hip OA confers robust neurovascular protection against AD, a profound biological antagonism that is clinically masked by depression-mediated phenotypic comorbidity. These findings propose an evolutionary trade-off model within the bone-brain axis, underscoring the urgency of active hip OA pain management to mitigate depressive symptoms and decelerate cognitive aging, while cautioning against the uncritical repurposing of anabolic inhibitors across these interconnected systems.

## Introduction

Osteoarthritis (OA) and Alzheimer’s disease (AD) represent two of the most prevalent and debilitating age-related conditions, collectively imposing an immense burden on global healthcare systems [1, 2]. Traditionally, these conditions have been conceptualized as anatomically distinct pathologies—one an age-related degeneration of peripheral joints, and the other a central neurodegenerative disorder. However, the emerging paradigm of the "bone-brain axis" has fundamentally shifted our understanding, revealing a profound and bidirectional neuro-endocrine-immune crosstalk between the skeletal system and the brain [3, 4]. Accumulating observational studies and large-scale cohort analyses have demonstrated that individuals with OA face a significantly increased risk of incident cognitive impairment and AD, whereas active pharmacological or surgical treatment of OA can attenuate this dementia risk [5, 6]. Currently, the prevailing consensus largely attributes this comorbidity to shared pathological drivers, positing that systemic low-grade inflammation, oxidative stress, and dysregulated autophagy act as common degenerative pathways bridging the joint and the brain [3, 7, 8].

Despite these epidemiological links, a striking conceptual contradiction remains when viewed through the lens of inverse comorbidity [9, 10]. Biologically, diseases driven by anabolic upregulation and abnormal cellular survival (such as the hypertrophic processes in OA) should theoretically antagonize diseases characterized by profound degeneration and apoptosis (such as AD). This raises a critical, unaddressed question: Is the observed increased AD risk in OA patients genuinely driven by shared systemic degeneration, or is it a phenotypic illusion confounded by secondary symptoms? Notably, chronic pain—the hallmark symptom of OA—has recently been conceptualized within a "brain aging framework," demonstrating that persistent nociceptive input and its associated psychological distress can actively accelerate biological brain aging [11]. The resulting "pain-depression" axis frequently manifests as cognitive decline, potentially masking the underlying genetic and biological reality of the bone-brain interplay [12]. Importantly, recent multidimensional analyses have established a profound bidirectional phenotypic and genetic architecture between skeletal vulnerabilities (such as fracture risk) and major depressive disorder [13]. This suggests that the psychological distress secondary to joint degeneration may be a critical, yet overlooked, confounder in the bone-brain axis.

While genomic tools like Mendelian randomization (MR) have been employed to disentangle such complex causal relationships, previous global MR studies utilizing standard organism-wide GWAS summary statistics are often confounded by horizontal pleiotropy and fail to capture the profound cellular heterogeneity of the brain [14, 15]. Specifically, the localized genetic interactions at the neurovascular unit (NVU) and the blood-brain barrier (BBB)—structures pivotal to AD pathogenesis—remain entirely unexplored in the context of systemic skeletal traits [16, 17].

Therefore, this study aims to resolve the "bone-brain paradox" by integrating longitudinal phenotypic data from the UK Biobank, advanced cell-type-stratified genomic modeling (MiXeR, MR), and validation through single-nucleus transcriptomics (snRNA-seq) from the ROSMAP cohort.

## Methods

### Observational Analysis (UK Biobank)

The study population was drawn from the full UK Biobank cohort (*N* = 502,359). After excluding participants of non-European ancestry, those with missing covariates, and individuals with prevalent dementia or insufficient follow-up (as detailed in Figure 1a), a total of 261,767 participants were included in the final longitudinal analysis. The UK Biobank study was approved by the North West Multi-centre Research Ethics Committee (REC reference: 11/NW/0382), and all participants provided written informed consent. This research was conducted using the UK Biobank Resource with accession ID 41376. Hip OA was defined using self-reports and ICD-10 hospital codes (M16), while incident AD was identified via ICD-10/EHR records (Additional file 1: Table S1). We constructed Cox proportional hazards models to estimate the Hazard Ratio (HR) of hip OA for incident AD. To account for the competing risk of death, we additionally employed Fine-Gray proportional subdistribution hazards models to estimate Subdistribution Hazard Ratios (SHR), treating all-cause mortality as a competing event. Models were adjusted for age, sex, BMI, education, income, smoking, alcohol intake, physical activity, history of depression, and *APOE* ε4 carrier status. We assessed the bidirectional relationship between phenotypic hip OA and incident AD. However, the reverse analysis (impact of baseline AD on incident hip OA) was not performed due to the low prevalence of AD at recruitment in the UK Biobank (*N* = 52), which precluded valid statistical modeling.

**Figure 1:**
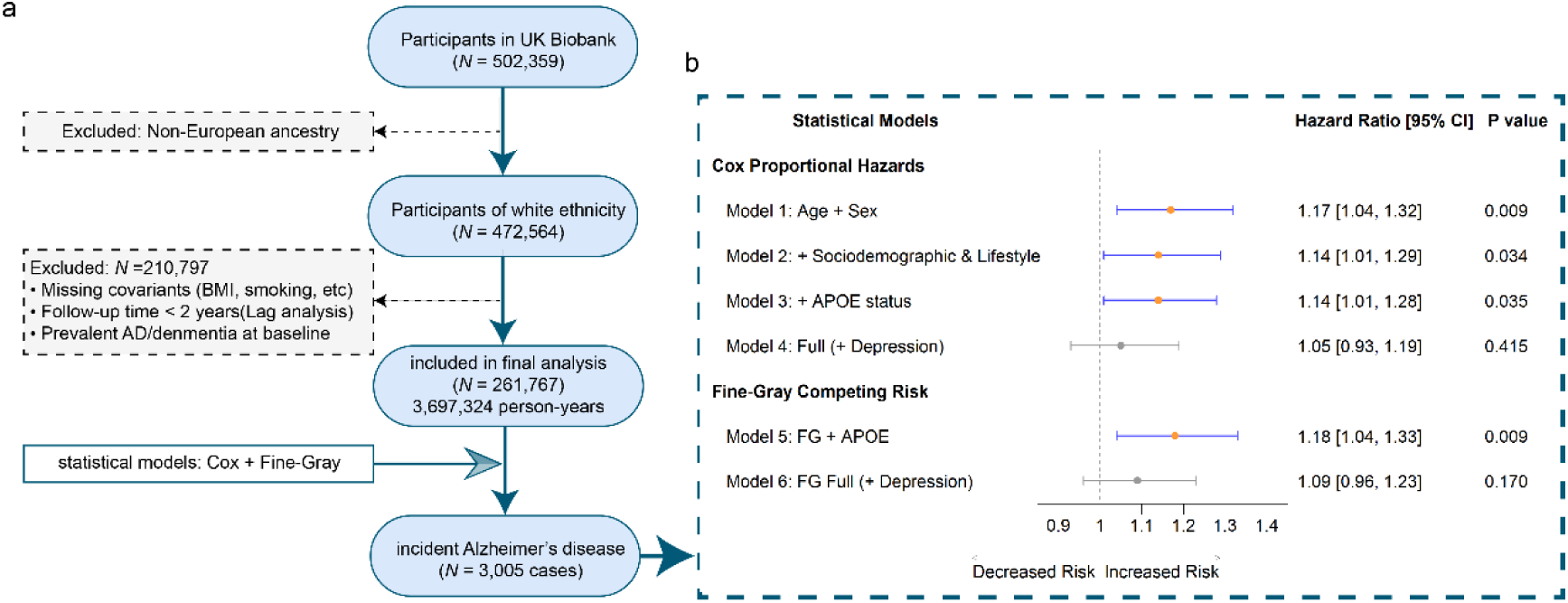
Study design and depression-mediated phenotypic association between hip osteoarthritis and Alzheimer’s disease. **(a)** Flowchart of participant selection in the UK Biobank. After excluding individuals of non-European ancestry, those with missing covariates, and individuals with prevalent dementia, 261,767 participants were included in the final analysis. The final cohort accumulated 3,697,324 person-years, with 3,005 incident AD cases identified. **(b)** Forest plot of the association between hip OA and incident AD using Cox proportional hazards (Models 1–4) and Fine-Gray competing risk models (Models 5–6). Points represent Hazard Ratios (HR) or Subdistribution Hazard Ratios (SHR) with 95% confidence intervals. Sequentially adjusted models demonstrate that in age- and sex-adjusted (Model 1) and sociodemographic- and lifestyle-adjusted analyses (Model 2; additionally adjusted for education, income, BMI, smoking, alcohol intake, and physical activity), hip OA was significantly associated with increased AD risk. This risk persisted after further adjusting for *APOE* ε4 carrier status (Model 3). However, the association was fully attenuated and became non-significant after adjusting for a history of depression (Model 4), indicating complete phenotypic mediation. This finding was corroborated by Fine-Gray models accounting for the competing risk of mortality, where the significant association in Model 5 disappeared upon the inclusion of depression in Model 6. Blue markers denote statistically significant associations, whereas grey markers denote non-significant findings. The vertical dashed line indicates the null value (1.0)

### Samples (GWAS Summary Statistics)

We utilized summary statistics from large-scale GWAS meta-analyses for hip osteoarthritis (hip OA; *N* = 393,873, *N*_case_ = 15,704) and Alzheimer’s disease (AD; *N* = 487,511, *N*_case_ = 39,106, *N*_proxy_ = 46,828). The hip OA data were derived from the UK Biobank and arcOGEN consortium [18], while the AD data were obtained from a meta-analysis of the European Alzheimer & Dementia Biobank (EADB) and UK Biobank proxy-AD phenotypes [19]. All studies were conducted in accordance with ethical guidelines, and informed consent was obtained from all participants or their proxies.

### Single-cell Expression Quantitative Trait Locus (sceQTL) Data

To dissect the cell-type-specific genetic architecture of brain-related traits, we utilized single-cell eQTL (sceQTL) summary statistics (derived from single-nucleus RNA sequencing) from Bryois et al. [20]. This dataset maps gene expression across eight major brain cell types (including astrocytes, endothelial cells, excitatory/inhibitory neurons, microglia, oligodendrocytes, oligodendrocyte precursor cells, and pericytes) in prefrontal and temporal cortical tissues from 192 individuals of European ancestry.

### MiXeR Analysis

We utilized MiXeR [21] to characterize the shared polygenic architecture, a robust approach recently validated in disentangling the complex pleiotropy between skeletal traits and neuropsychiatric disorders [13]. First, univariate MiXeR models were fitted to estimate each trait’s polygenicity (i.e., the number of causal variants explaining 90% of SNP heritability) and discoverability (i.e., the average effect size of detectable causal variants). We confirmed adequate model fit by verifying positive Akaike Information Criterion (AIC) and Bayesian Information Criterion (BIC) values, indicating sufficient statistical power for subsequent bivariate analyses (Additional file 1: Table S2). Next, bivariate MiXeR models were used to estimate the number of causal variants shared between each trait pair, as well as those unique to each trait. The models also provided estimates of the proportion of shared causal variants with concordant effect directions. Polygenic overlap was further quantified using the Dice coefficient (Additional file 1: Table S3). Conditional quantile–quantile (Q–Q) plots were generated to visualize cross-trait enrichment, stratifying *P*-values of the primary trait by association strength with the secondary trait at three significance thresholds (*P* ≤ 0.1, 0.01, and 0.001).

### Two-Sample Mendelian Randomization (MR) Analysis

To validate the genetic correlations identified by MiXeR, we performed two-sample Mendelian randomization (MR) analyses for hip OA–AD associations. The same GWAS summary statistics were used as in the MiXeR analyses. To strictly ensure the independence of instrumental variables (IVs), linkage disequilibrium (LD) clumping was performed (window size: 250 kb, *r*² < 0.1) based on the European 1000 Genomes reference panel. We employed multiple MR methods to improve robustness: the IVW method, the simple median estimator, and MR-PRESSO to detect and correct for horizontal pleiotropy [14].

### Cell-Type-Specific Mendelian Randomization (csMR) Analysis

To fine-map potential causal effects of hip OA on AD across distinct brain cell types, we applied cell-type-specific Mendelian randomization (csMR) [22]. This approach leverages sceQTL data to prioritize causal genes and cell types underlying cross-trait associations. We used the same GWAS summary statistics for hip OA and AD, along with sceQTL data from the eight brain cell subtypes described above. Variants within the major histocompatibility complex (MHC) region (chr6: 28,477,797–33,448,354; GRCh37) were excluded due to high LD and complex haplotype structure. Credible sets of causal variants were defined with a posterior probability threshold of 0.9. Colocalization between GWAS signals and eQTLs was assessed within ± 100 kb of gene transcription start sites using default parameters. Strong evidence of colocalization was defined as a posterior probability of the fourth hypothesis (PPH4) > 0.8. For instrumental variable selection, we first identified SNPs associated with hip OA at genome-wide significance (*P* < 5 × 10^-8^). LD clumping was performed using PLINK (window size: 250 kb, *r*^2^ < 0.1), yielding a set of index SNPs. These were then intersected with outcome datasets; for missing SNPs, high-quality proxies (*r*^2^ > 0.8 in European 1000 Genomes reference panel) were used. To strictly ensure instrumental validity, particularly for loci restricted to a single instrument (*N*_IV_ = 1), only variants showing strong evidence of colocalization (PPH4 > 0.8) were retained for the final analysis. To account for multiple testing across the eight brain cell types, we applied a Bonferroni correction. Results with *P* < 0.00625 (0.05 / 8) were considered statistically significant, while *P* values between 0.00625 and 0.05 were considered nominally significant (Additional file 1: Table S6).

### Conjunctional/Conditional False Discovery Rate (ccFDR) Analysis

To identify specific pleiotropic genomic loci jointly associated with hip OA and AD—independent of global genetic correlation directionality—we applied conjunctional false discovery rate (conjFDR) analysis [23, 24]. This empirical Bayesian statistical framework robustly identifies shared loci by conditioning the test statistics of the primary trait on the significance of the secondary trait, and vice versa. Genomic loci satisfying a conservative threshold of conjFDR < 0.05 were deemed to have a statistically significant shared polygenic overlap.

### FUMA and Metascape Functional Annotation

Genomic loci satisfying a conservative threshold of conjFDR < 0.05 were submitted to Functional Mapping and Annotation (FUMA) v1.5.2 [25] for functional annotation and genomic mapping. Independent significant SNPs were identified using LD-based clumping (distance ≤ 250 kb, *r*^2^ < 0.6) with the European 1000 Genomes reference panel. Lead SNPs within each locus were defined using a stricter LD threshold (*r*^2^ < 0.1) and selected based on the lowest conjFDR value (Additional file 1: Table S7). Each locus was annotated to genes based on physical position (within gene body) or proximity to the nearest transcription start site. We assessed the novelty of lead SNPs by evaluating whether they reached genome-wide significance (*P* < 5 × 10^-8^) in the original hip OA or AD GWAS. Functional pathway enrichment analysis of the shared risk genes was performed using Metascape [26].

### Single-nucleus RNA-sequencing (snRNA-seq) Transcriptomic Validation

To validate the cell-type-specific causal inferences derived from the Mendelian randomization analyses within a biological context, we queried human brain single-nucleus RNA-sequencing (snRNA-seq) data from the Religious Orders Study and Memory and Aging Project (ROSMAP) cohort. Clinical metadata and processed count matrices were obtained to assess the basal transcriptional landscape and disease-associated dynamics of prioritized shared risk genes (*MAPT*, *MAPT-AS1*, *PIK3CA*, *AKT1*, *AKT2*, and *AKT3*) within the neurovascular unit and glial support network. To ensure precise phenotypic stratification, clinical data were processed by extracting patient identifiers (projid) and final consensus cognitive diagnosis scores (cogdx). Participants were strictly categorized into two primary analytical groups based on their clinical cognitive status at the time of death: cognitively unimpaired individuals ("Control", defined as cogdx = 1) and patients with a definitive clinical diagnosis of Alzheimer’s dementia ("AD", defined as cogdx = 4 or 5). Individuals with intermediate cognitive impairments, such as mild cognitive impairment (MCI), or other neurological conditions, were categorized as "Other" and excluded from the primary differential expression visualizations to maximize the signal-to-noise ratio.

The processed count matrices and stratified clinical metadata were integrated to construct Seurat objects utilizing the Seurat R package (version 4.0+). To account for sequencing depth variations, raw gene counts were log-normalized and scaled prior to downstream analysis. Cells were then clustered and annotated into five previously defined target brain cell types: Astrocytes, Endothelial cells, Inhibitory neurons, Oligodendrocyte precursor cells, and Pericytes. Cell populations were clustered into five previously annotated target brain cell types: Astrocytes, Endothelial cells, Inhibitory neurons, Oligodendrocyte precursor cells, and Pericytes. Basal gene expression profiles across these cell populations were visualized using dot plots, representing both the proportion of expressing cells and average scaled expression. To evaluate disease-associated transcriptional remodeling, split violin plots were generated to compare the expression distribution of the target genes between the defined Control and AD groups within each specific cell type. All single-cell data processing and visualization steps were performed using the Seurat package in R.

### Statistical Analysis

All statistical analyses were conducted using R software (version 4.2.1). Observational survival analyses, including Cox proportional hazards and Fine-Gray competing risk models, were performed using the survival and cmprsk packages. Genomic and MR analyses were implemented utilizing the TwoSampleMR, MendelianRandomization, and MR-PRESSO packages. The processed count matrices and stratified clinical metadata were integrated to construct Seurat objects utilizing the Seurat R package (version 4.0+) [27]. Unless otherwise specified with a stringent multiple-testing correction (e.g., Bonferroni correction in the csMR analysis), a two-sided *P* value < 0.05 was considered statistically significant.

## Results

### Depression mediates the observational association between hip osteoarthritis and Alzheimer’s disease

We included 261,767 participants of white ethnicity in the final phenotypic analysis (Figure 1a). During a median follow-up of 14.46 years (interquartile range [IQR]: 13.73–15.17 years), accumulating 3,697,324 person-years, we identified 3,005 incident cases of Alzheimer’s disease (AD). In the age- and sex-adjusted model, individuals with hip OA exhibited a significantly elevated risk of developing AD compared to controls (Hazard Ratio [HR] 1.17; 95% CI 1.04–1.32; *P* = 0.009). This association remained robust after further adjustment for sociodemographic (education, income) and lifestyle factors (BMI, smoking, alcohol consumption, physical activity) (HR 1.14; 95% CI 1.01–1.29; *P* = 0.035). Notably, additional adjustment for the *APOE* ε4 genotype did not alter the risk estimates (HR = 1.14; 95% CI 1.01–1.28; *P* = 0.035), suggesting that the observed phenotypic association is not driven by *APOE*-related genetic pleiotropy (Figure 1b).

However, the association between hip OA and AD was fully attenuated upon introducing depression into the multivariable model (HR = 1.05; 95% CI 0.93–1.19; *P* = 0.415). This mediating effect implies that the "pain-depression" axis may account for the observed comorbidity. Sensitivity analyses accounting for the competing risk of death corroborated these findings; the subdistribution hazard ratio (SHR) was significant in models excluding depression (SHR = 1.18; 95% CI 1.04–1.33; *P* = 0.009) but became non-significant after adjustment for depression (SHR = 1.09; 95% CI 0.96–1.23; *P* = 0.17) (Figure 1b).

### Genomic architecture reveals antagonistic polygenic overlap between hip osteoarthritis and Alzheimer’s disease

Evaluation of univariate model fit statistics (Akaike and Bayesian Information Criteria; AIC and BIC) confirmed sufficient statistical power for bivariate MiXeR modeling (Additional file 1: Table S2 and 3). By comparing model fits, we determined that the specific MiXeR-estimated architecture provided a significantly better fit than the minimal overlap hypothesis (AIC = 11.28, BIC = 1.72). The estimated model also outperformed the maximal overlap hypothesis based on AIC (9.20), though the distinction was less pronounced under the more conservative BIC threshold (-0.37).

Despite a modest global genetic correlation (*r_g_* = -0.11, SE = 0.02) and a Dice coefficient of 6.82% indicating limited total polygenic overlap (Figure 2a), the internal structure of the shared genetics revealed a strong inverse relationship. We identified 3,223 and 218 putatively causal variants associated with hip OA and AD, respectively, with an intersection of 116 shared variants. Crucially, the correlation of effect sizes within this shared component was strongly negative (ρβ = -0.84, SD = 0.17). Specifically, 83.7% of these shared variants exhibited discordant effect directions—where risk-increasing alleles for one trait exert protective effects on the other—while only 16.3% showed consistent directionality. This significant polygenic overlap was further corroborated by conditional quantile–quantile (Q–Q) plots, which demonstrated that SNP enrichment for hip OA scaled progressively with the strength of their association with AD, and vice versa (Figure 2b and 2c).

**Figure 2:**
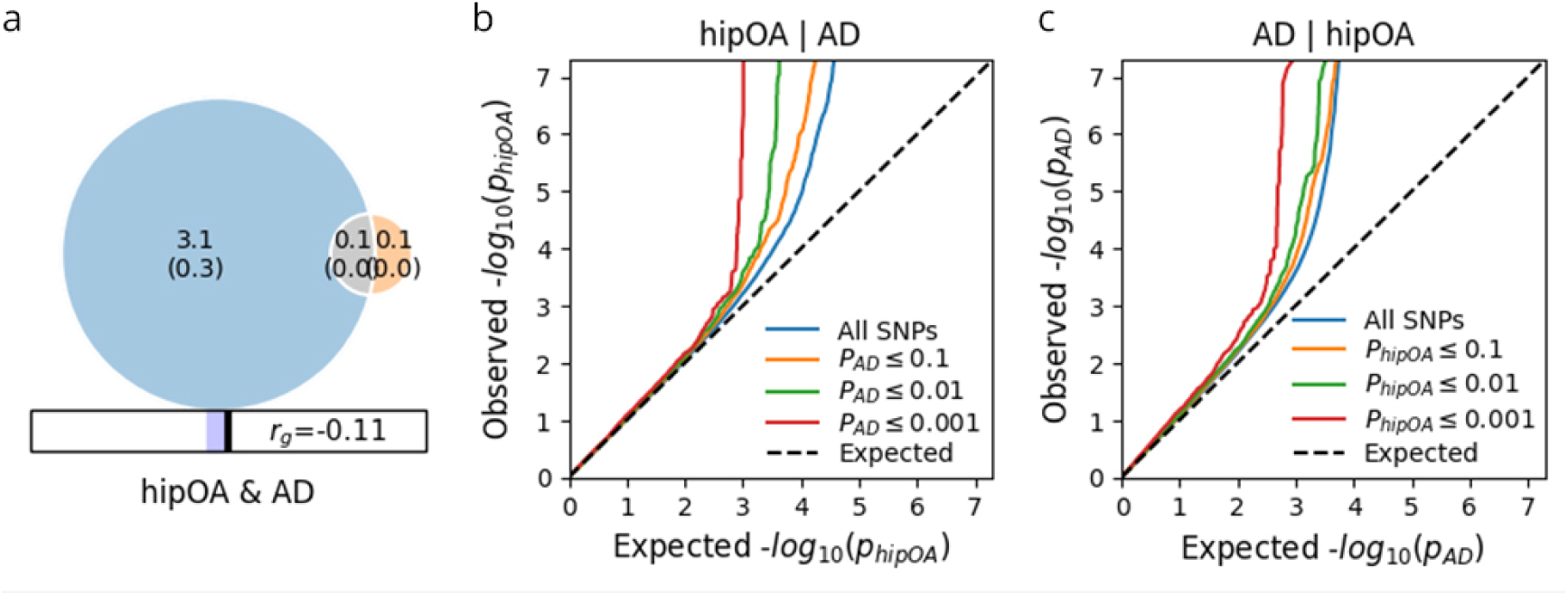
Polygenic architecture and shared genetic basis between hip osteoarthritis and Alzheimer’s disease. **(a)** MiXeR graphical representation of the polygenic overlap. The Venn diagram visualizes the estimated number of causal variants (in thousands) for hip osteoarthritis (blue) and Alzheimer’s disease (orange), alongside their shared component (grey). The numbers indicate the estimated count of causal variants (e.g., 3.1 represents ∼3,100 variants) with standard errors in parentheses. The intersection suggests approximately 116 shared variants influencing both traits. The color bar below denotes the genetic correlation (*r_g_* = -0.11) within the overlapping polygenic component, indicating a negative correlation of effect sizes. **(b, c)** Conditional quantile–quantile (Q–Q) plots demonstrating polygenic enrichment. **(b)** Empirical distribution of nominal -log_10_ (*P*) values for hip OA SNPs, conditioned on their significance in the AD GWAS at increasingly stringent thresholds (orange: *P* ≤ 0.1; green: *P* ≤ 0.01; red: *P* ≤ 0.001). The successive leftward deflection of the curves from the null expectation (dashed black line) indicates that variants associated with AD are significantly enriched for hip OA susceptibility. **(c)** Reciprocal analysis showing the distribution of AD *P*-values conditioned on their association with hip OA. The robust stratification confirms a bidirectional polygenic overlap between the two phenotypes.

### Mendelian randomization reveals asymmetric causal protection between Alzheimer’s disease and hip osteoarthritis

Following data harmonization to exclude ambiguous palindromic SNPs and missing variants, we identified 51 independent SNPs as valid instrumental variables (IVs) to investigate the causal effect of hip OA on AD. The *F*-statistics for these IVs ranged from 29.74 to 105.20 (mean = 39.51), indicating strong genetic validity and the absence of weak instrument bias (Additional file 1: Table S4). The primary inverse-variance weighted (IVW) analysis initially suggested a nominal causal association, where genetically predicted hip OA was associated with a reduced risk of AD (OR = 0.929, *P* = 0.009). However, we observed significant heterogeneity across the estimates (Cochran’s *Q* = 179.7, *P* < 0.001). Furthermore, the MR-PRESSO Global test detected significant horizontal pleiotropy (*P* < 0.001). Sensitivity analyses using the Weighted Median method (OR = 0.984, *P* = 0.525) and MR-Egger regression (OR = 1.006, *P* = 0.936) did not support a significant causal relationship. The MR-Egger intercept term was -0.008 (*P* = 0.254), suggesting no statistical evidence of directional pleiotropy from the Egger perspective, although the PRESSO test was more sensitive. Crucially, the MR-PRESSO outlier test identified four specific pleiotropic SNPs: rs199529, rs35489312, rs55938136, and rs62063281. After removing these outliers, the causal estimate from the IVW method was substantially attenuated and no longer statistically significant (Outlier-corrected β = -0.034, equivalent to OR = 0.967; *P* = 0.126). The MR-PRESSO Distortion test confirmed a significant difference between the estimates before and after outlier removal (*P* = 0.019). Collectively, these findings indicate that the initial significant association between hip OA and AD risk was likely driven by horizontal pleiotropy rather than a robust causal effect.

Similarly, applying identical harmonization criteria, we identified 186 independent SNPs as valid IVs to investigate the reverse causal effect of AD liability on hip OA. The instruments exhibited strong genetic validity, with *F*-statistics ranging from 29.73 to 519.11 (mean = 84.08), indicating no evidence of weak instrument bias (Additional file 1: Table S5).

Genetically predicted AD was associated with a reduced risk of hip OA (IVW OR = 0.962, 95% CI: 0.938–0.986, *P* = 0.002). Although heterogeneity and horizontal pleiotropy were detected, the protective effect remained robust after correcting for outliers (MR-PRESSO corrected OR = 0.965, *P* = 0.003). Sensitivity analyses using Weighted Median and MR-Egger methods showed directionally consistent estimates, further supporting a causal protective effect of AD liability on hip OA (Figure 3).

**Figure 3:**
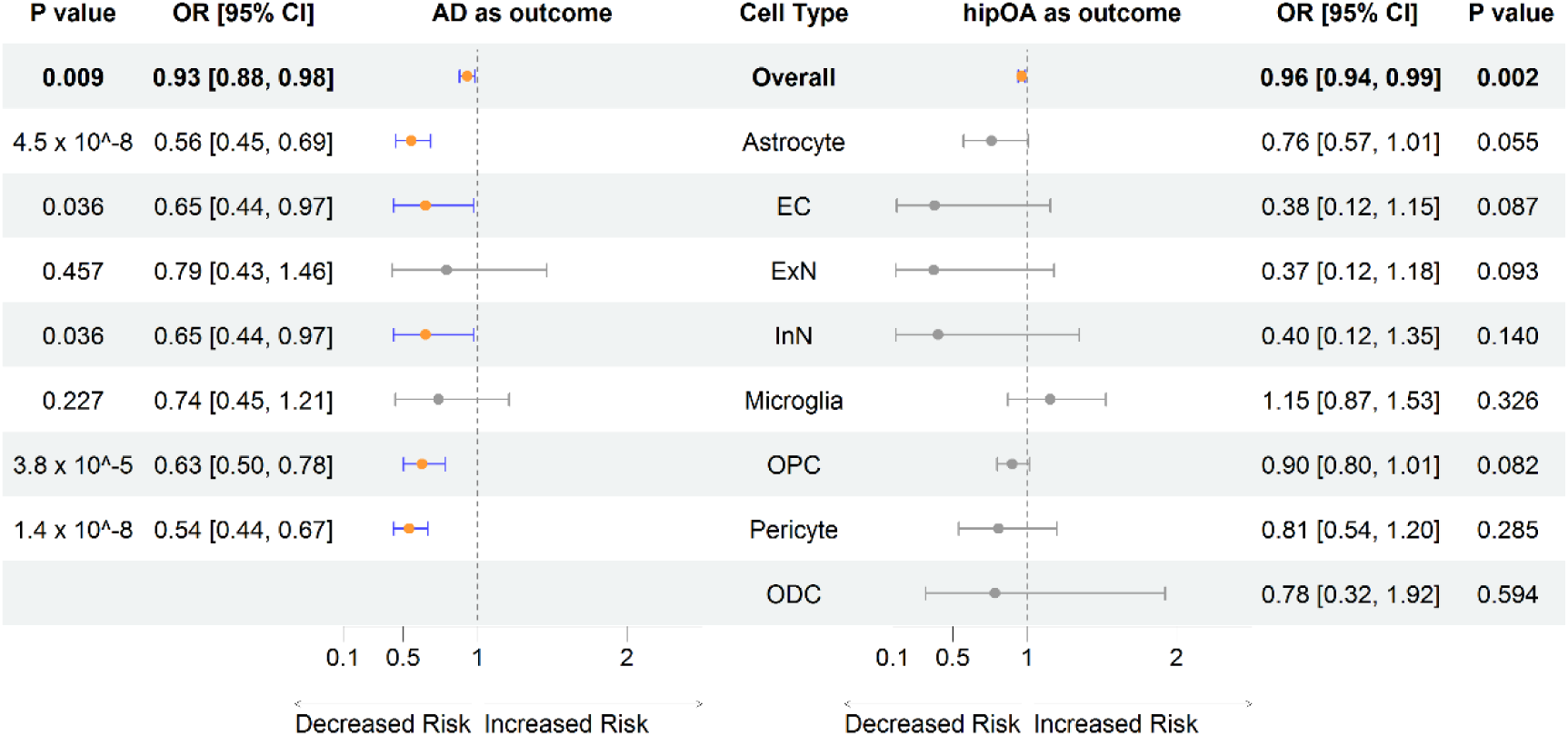
Bidirectional and Cell-Type-Specific Causal Associations Between Hip Osteoarthritis and Alzheimer’s Disease. The forest plot is organized into two panels based on the outcome phenotype. The left panel displays the causal effect of genetic liability to hip osteoarthritis (hip OA) on Alzheimer’s disease (AD) risk. The top row represents the global inverse-variance weighted (IVW) estimate (*P* = 0.009). The rows below show cell-type-stratified MR (csMR) results, identifying robust, Bonferroni-significant protective effects in Pericytes (*P* = 1.4 × 10^-8^), Astrocytes (*P* = 4.5 × 10^-8^), and Oligodendrocyte Precursor Cells (OPC; *P* = 3.8 × 10^-5^). The bottom row for Oligodendrocytes (ODC) contains no data because no valid instrumental variables were identified (*N*_IV_ = 0) for this cell type. The right panel presents the reverse analysis (AD liability on hip OA risk). While the global analysis (top row) indicates a significant protective effect (*P* = 0.002), the cell-type-specific estimates (lower rows) are generally attenuated or non-significant, suggesting the protection is systemic rather than cell-specific. Data are presented as Odds Ratios (OR) with 95% confidence intervals. *P* values are derived from IVW (for *N*_IV_ ≥ 2) or Wald ratio (for *N*_IV_ = 1) methods. Abbreviations: EC, endothelial cells; ExN, excitatory neurons; InN, inhibitory neurons; ODC, oligodendrocytes; OPC, oligodendrocyte precursor cells.

### Cell-type-specific Mendelian randomization identifies neurovascular and glial drivers of the protective effect

Given that standard global MR estimates lack cellular resolution and were likely confounded by horizontal pleiotropy, we next sought to determine if the true causal signals were specific to distinct cellular contexts. To test this, we performed cell-type-stratified Mendelian randomization (csMR) across eight brain cell types. To ensure instrumental validity, we restricted our analysis to variants with strong evidence of colocalization. This stringent filtering revealed a striking cell-type-specific protective profile (Additional file 1: Table S6).

This analysis revealed a striking cell-type-specific protective profile. We observed robust, Bonferroni-significant evidence that genetic liability to hip OA confers protection against AD in pericytes (*N*_IV_ =1; OR = 0.540; 95% CI 0.437–0.668; *P* =1.40 × 10^-8^), astrocytes (*N*_IV_ =1; OR = 0.556; 95% CI 0.451–0.686; *P* = 4.50 × 10^-8^), and oligodendrocyte precursor cells (OPCs; *N*_IV_ =3; OR = 0.628; 95% CI 0.503–0.783; *P* = 3.76 × 10^-5^). Nominally significant protective associations were identified in endothelial cells and inhibitory neurons (both OR ≈ 0.65; *P* = 0.036). Notably, these identical estimates arose because the stringent colocalization filtering identified the same set of instrumental variables (rs2267847 and rs974295) as valid causal drivers in both cell types, reflecting a shared cis-regulatory architecture at these specific loci. In contrast, no causal estimate for the effect of hip OA on AD could be generated for oligodendrocytes (ODC) due to the absence of instrumental variables capable of passing the strict colocalization threshold (PPH4 > 0.8). The Wald ratio method was employed for cell types with a single instrument, while the inverse-variance weighted (IVW) method was used for those with multiple instruments. Collectively, these results implicate the neurovascular unit (pericytes, endothelial cells) and glial support (astrocytes, OPCs) as the primary cellular mediators of the inverse comorbidity.

### Shared genomic architecture is driven by antagonism at the MAPT locus and PI3K/AKT signaling

To pinpoint specific genomic regions driving this inverse relationship, we utilized conditional false discovery rate (conjFDR) analysis. At a threshold of conjFDR < 0.05, we identified 3,000 pleiotropic loci associated with both phenotypes (Figure 4a). Consistent with the global correlation results, the vast majority of these shared lead SNPs (92.8%) exhibited discordant effect directions, confirming that the genetic architecture is dominated by biological antagonism.

**Figure 4:**
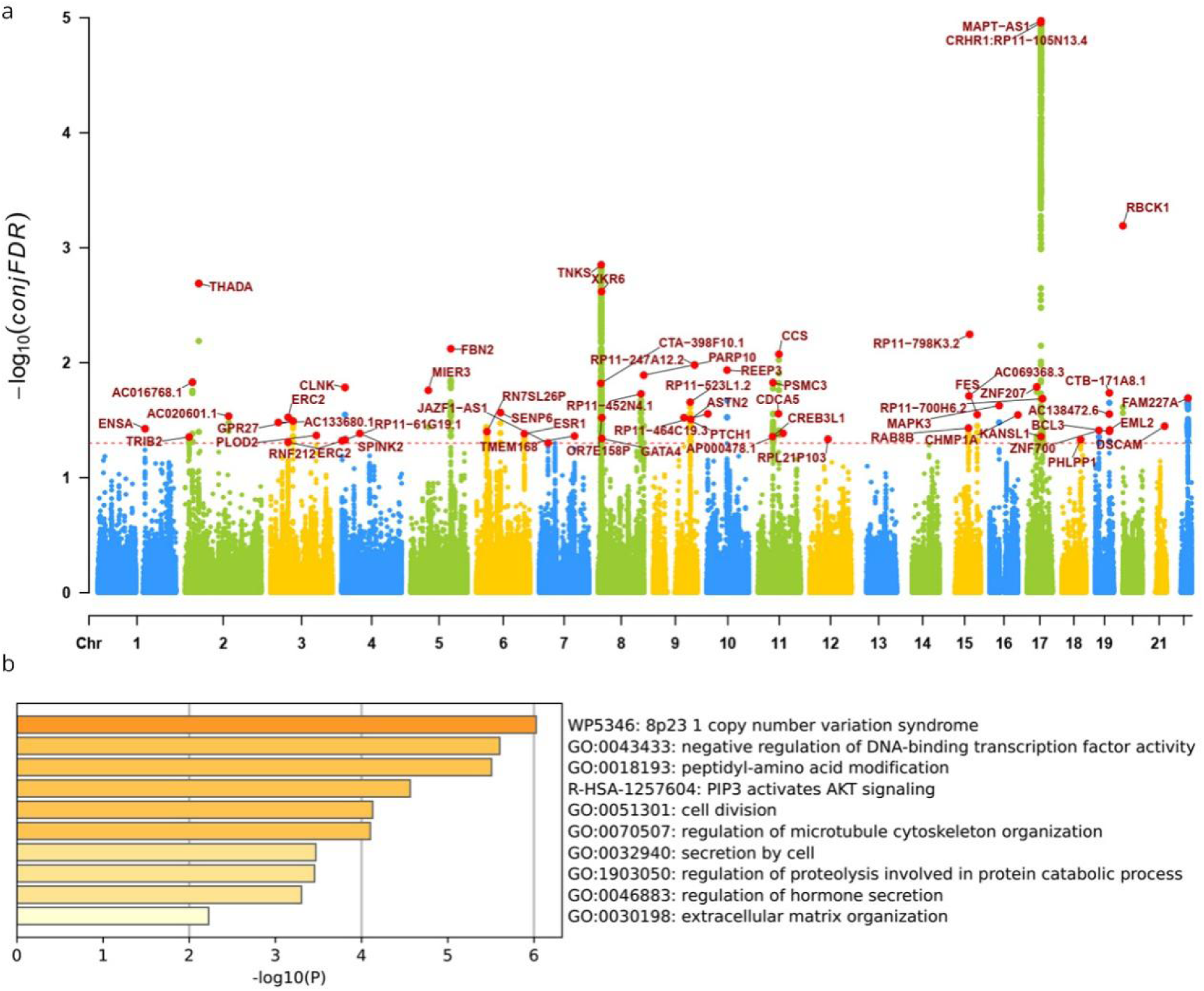
Identification of pleiotropic loci and shared biological pathways between hip osteoarthritis and Alzheimer’s disease. **(a)** Conditional Manhattan plot displaying the specific genetic loci shared between hip osteoarthritis (hip OA) and Alzheimer’s disease (AD). The y-axis represents the conditional false discovery rate (-log_10_(conjFDR)) for hip OA given the association with AD (and vice versa). Significant loci (conjFDR < 0.05) are highlighted above the red dashed significance threshold line. Notable peaks include the *MAPT-AS1* locus on chromosome 17, which exhibits the strongest shared association signals. Independent lead SNPs are annotated with their nearest gene names. **(b)** Functional pathway enrichment analysis of the shared risk genes identified in (a), performed using Metascape. The bar chart ranks the top enriched ontology terms and pathways by statistical significance (-log_10_(*P*)). Key enriched biological processes include "PIP3 activates AKT signaling" and "regulation of microtubule cytoskeleton organization," supporting a shared mechanistic basis involving anabolic signaling and cytoskeletal stability.

We further refined these signals into independent causal loci using strict linkage disequilibrium clumping (*r^2^* < 0.1, distance ≤ 250 kb), identifying 60 distinct lead SNPs (Additional file 1: Table S7). Among the top nine most significant hits, all displayed opposite effects on hip OA and AD. The strongest signal mapped to the 17q21.31 region. Specifically, the lead variant rs62055517 resides within the intron of *MAPT-AS1* (*MAPT* Antisense RNA 1), a long non-coding RNA known to regulate *MAPT* expression, while the secondary hit rs55938136 is located upstream of the *MAPT* gene itself. Functional enrichment analysis (Metascape) of genes mapped to these 60 independent lead SNPs highlighted "PIP3 activates AKT signaling" and "Regulation of microtubule cytoskeleton organization" as top biological processes. These findings suggest a mechanistic model where shared genetic variants modulate PI3K/AKT signaling and cytoskeletal stability to exert opposing effects—driving anabolic hypertrophy in the joint while conferring neuroprotective resilience in the brain.

### Single-nucleus transcriptomic validation reveals disease-associated remodeling of the PI3K/AKT and MAPT axes in the neurovascular unit and glia

To contextualize our cell-type-specific Mendelian randomization findings within a pathologically relevant biological framework, we queried single-nucleus RNA sequencing (snRNA-seq) data from the ROSMAP cohort. Specifically, we sought to map the basal expression and disease-associated transcriptional dynamics of our prioritized shared targets (*MAPT*, *PIK3CA*, and the *AKT* family) across the implicated neurovascular unit (pericytes, endothelial cells [EC]) and glial support network (astrocytes, oligodendrocyte precursor cells [OPC], and inhibitory neurons [InN]). As expected, while transcripts with lower nuclear abundance or technical capture limitations in snRNA-seq (such as the lncRNA *MAPT-AS1*, *AKT1*, and *AKT2*) exhibited characteristic sparsity, the core mechanistic drivers demonstrated robust expression profiles (Figure 5a). Notably, *MAPT* exhibited profound expression not only in InN but also robustly localized within astrocytes and OPCs. Concurrently, the upstream PI3K catalytic subunit (*PIK3CA*) and the downstream neuroprotective effector *AKT3* were pervasively expressed across all five cell types, validating that these specific cellular populations possess the necessary transcriptional machinery to mediate the shared genetic signals.

**Figure 5.**
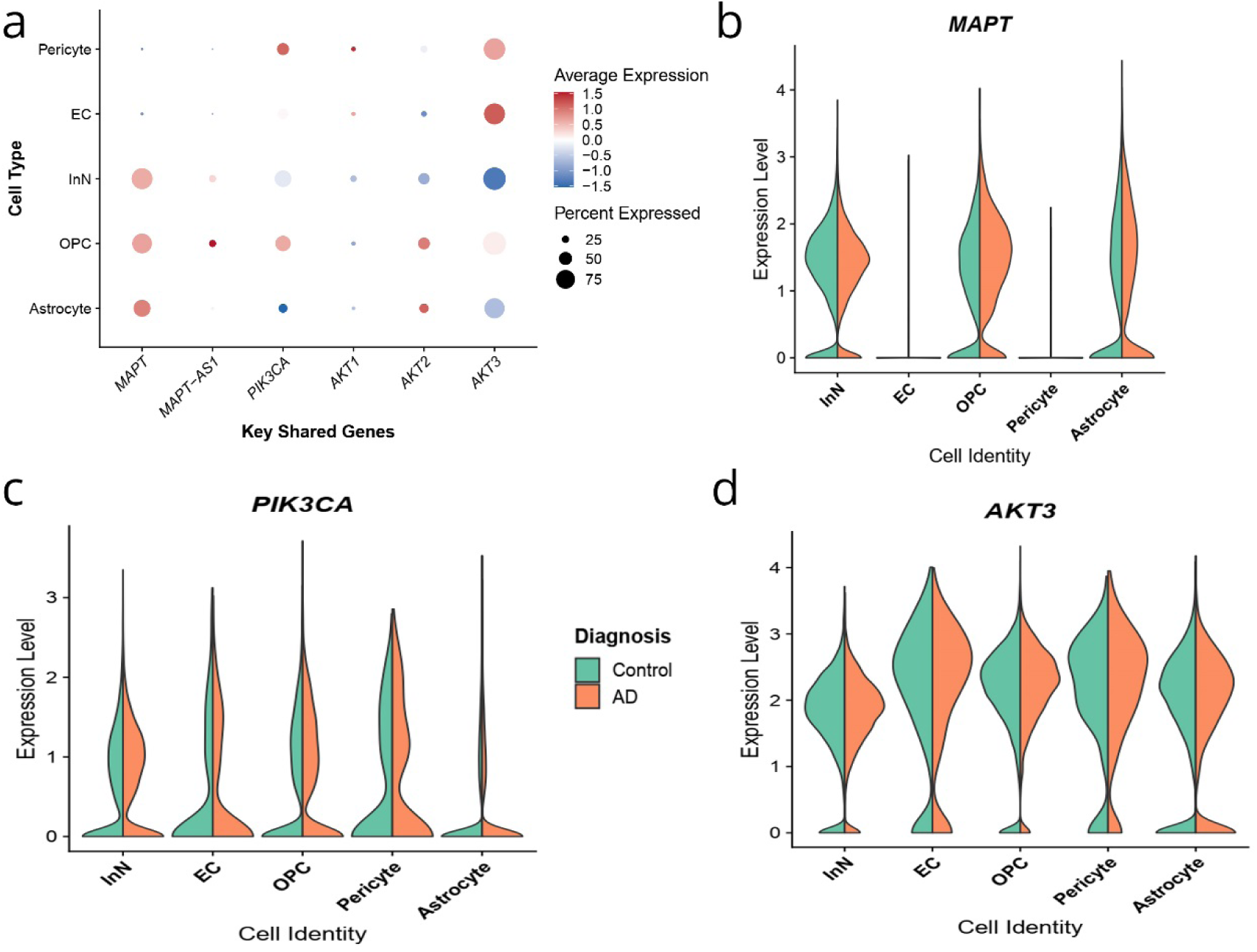
Single-nucleus transcriptomic validation of shared genetic drivers within the neurovascular and glial milieu. Utilizing single-nucleus RNA sequencing (snRNA-seq) data from the ROSMAP cohort, this figure validates the cellular localization and disease-associated transcriptional dynamics of key genes implicated by genetic modeling. **(a)** Dot plot illustrating the basal transcriptional landscape of prioritized shared risk genes across the five critical brain cell populations identified via cell-type-stratified Mendelian randomization: Astrocytes, Oligodendrocyte precursor cells (OPC), Inhibitory neurons (InN), Endothelial cells (EC), and Pericytes. Dot size is proportional to the percentage of cells expressing the target gene within each cluster, while the color gradient represents the scaled average expression level (blue = lower, red = higher relative expression). Note the robust cellular ubiquity of the core signaling components *PIK3CA* and *AKT3*, alongside the distinct localization of *MAPT* in glial and neuronal subtypes. This contrasts with the technical sparsity characteristic of low-abundance transcripts (*MAPT-AS1*, *AKT1*, *AKT2*) typically observed in snRNA-seq. **(b–d)** Split violin plots illustrating disease-associated transcriptional remodeling of the robustly expressed core effector genes. The expression density distributions of **(b)** *MAPT*, **(c)** *PIK3CA*, and **(d)** *AKT3* are compared side-by-side between cognitively unimpaired controls (Green, left distribution) and patients with clinical Alzheimer’s disease (Orange, right distribution) within each cell type. This visualization highlights context-specific distributional shifts and dynamic modulation of the *MAPT* and PI3K/AKT signaling axes in AD pathology, particularly within the neurovascular unit and supporting glial networks.

Focusing our subsequent disease-state analysis on these robustly expressed core effectors, stratification of these cell populations by clinical diagnosis (Control vs. Alzheimer’s disease) revealed distinct disease-associated transcriptional remodeling (Figure 5b–d). The expression density distributions of *MAPT* within astrocytes and OPCs, as well as *AKT3* within the neurovascular unit (pericytes and ECs), exhibited observable topological shifts in AD patients compared to cognitively unimpaired controls (Figure 5b and 5d). Collectively, these human-derived transcriptomic data substantiate our causal genetic inferences, confirming that the key drivers of the protective antagonistic pleiotropy are actively transcribed and pathologically responsive within the specific neurovascular and glial contexts.

## Discussion

The present study resolves the profound discordance between the observational comorbidity of hip OA and AD and their underlying biological antagonism. By integrating multi-omic architectures with longitudinal phenotypic analysis, we propose a comprehensive evolutionary trade-off model that conceptualizes the "bone-brain paradox" through the lens of the bone-brain axis [3, 28] (Figure 6).

**Figure 6:**
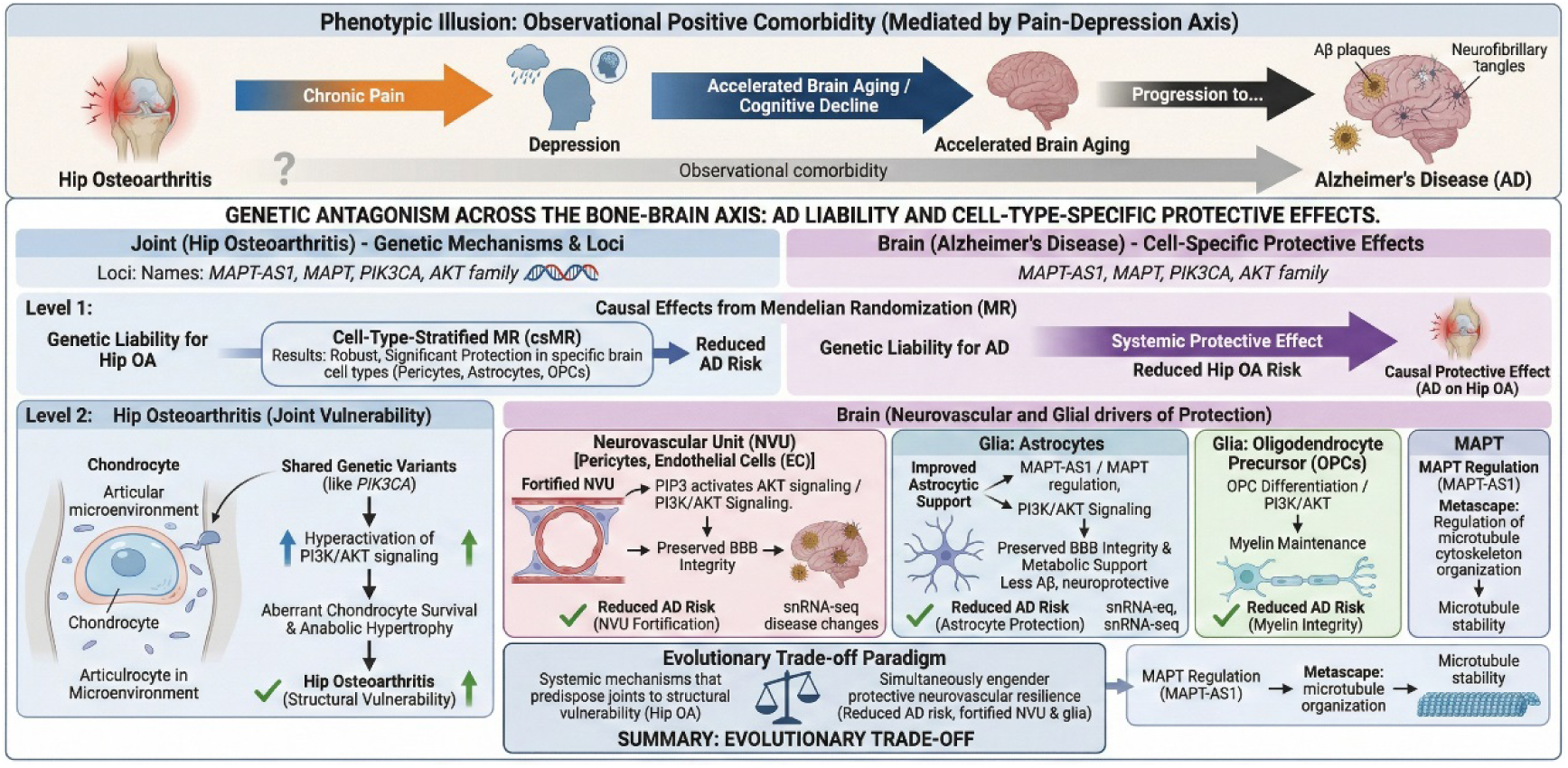
Mechanistic Paradigm of the Bone-Brain Axis: Phenotypic Illusion, Antagonistic Pleiotropy, and Evolutionary Trade-offs Linking Hip Osteoarthritis and Alzheimer’s Disease. **(a)** The Phenotypic Illusion (Upper Panel): The clinically observed positive comorbidity between hip osteoarthritis (hip OA) and Alzheimer’s disease (AD) is predominantly driven by the pain-depression axis rather than shared neurodegenerative pathology. Structural degeneration in hip OA induces persistent somatic pain, which catalyzes a depressive cascade. This chronic psychological stress and secondary depression deplete cognitive reserves and accelerate biological brain aging, ultimately propelling clinical AD progression. **(b)** Genetic Antagonism and Shared Architecture (Lower Panel, Level 1): Beneath the observational overlap lies a profound genetic antagonism. Global Mendelian randomization (MR) demonstrates that genetic liability to AD exerts a systemic protective causal effect against hip OA risk. Conversely, while global MR of hip OA on AD is confounded by horizontal pleiotropy, cell-type-stratified MR (csMR) reveals robust, causal neuroprotective effects localized specifically to the neurovascular unit (NVU) and supporting glial networks. **(c)** Cell-Type-Specific Mechanisms and Evolutionary Trade-off (Lower Panel, Level 2): Shared genetic loci (including *MAPT-AS1*, *MAPT*, *PIK3CA*, and the *AKT* family) exert highly tissue-specific antagonistic pleiotropy. In the articular microenvironment, shared variants drive hyperactivation of PI3K/AKT signaling, promoting aberrant chondrocyte survival and anabolic hypertrophy, which predisposes the joint to structural vulnerability (Hip OA). In stark contrast, within the brain, this identical anabolic drive and microtubule regulation (via MAPT) translate into a fortified neurovascular interface (pericytes and endothelial cells) and enhanced glial support (astrocytes and oligodendrocyte precursor cells [OPCs]). This fortification preserves blood-brain barrier (BBB) integrity and promotes myelin maintenance, shielding neural tissues from AD pathology. Collectively, these findings propose an evolutionary trade-off paradigm: systemic mechanisms that predispose joints to structural failure simultaneously engender protective neurovascular resilience in the aging brain.

Our findings initially reveal that the clinically observed hip OA-AD comorbidity is, in fact, a "phenotypic illusion" mediated predominantly by the pain-depression axis. While recent mechanistic reviews have heavily emphasized systemic inflammation and oxidative stress as the shared degenerative pathways linking these two conditions [3, 7, 8], our longitudinal survival analyses demonstrate that adjusting for a history of depression fully attenuates the positive association between hip OA and AD. This aligns perfectly with the recently proposed "brain aging framework" for chronic pain, which posits that persistent somatic pain depletes cognitive reserves and accelerates biological brain aging via chronic neuroinflammation and psychological stress [11, 29]. Chronic pain acts as a potent driver of depression, which intrinsically catalyzes cognitive decline [30-32]. This aligns with our recent findings demonstrating a robust bidirectional phenotypic and pleiotropic genetic link between skeletal integrity (e.g., fractures) and major depressive disorder [13]. Therefore, the structural degeneration in hip OA may not directly incite AD pathology; rather, it propels a depressive cascade that accelerates clinical cognitive decline. This elegantly contextualizes recent large-scale findings where active OA treatment was associated with a decreased risk of incident dementia [6, 33]—effective OA management likely mitigates peripheral nociception and secondary depression, thereby interrupting the pain-induced acceleration of brain aging rather than altering a shared neurodegenerative pathology [34, 35].

In stark contrast to this phenotypic illusion, our genomic analyses uncover a profound biological antagonism embedded within the bone-brain axis. While global MR signals were obscured by horizontal pleiotropy [14, 36], our cell-type-stratified approach revealed robust neuroprotective effects specifically localized to the NVU, primarily driven by pericytes and astrocytes. This indicates that the genetic variants mediating hip OA liability do not merely propagate systemic inflammation [7], but instead exert highly tissue-specific antagonistic pleiotropy. Mechanistically, our fine-mapping and pathway enrichment analyses converged on the *MAPT* locus and PI3K/AKT signaling. In the articular microenvironment, hyperactivation of PI3K/AKT drives aberrant chondrocyte survival and anabolic hypertrophy [37-39]; conversely, in the brain, this identical anabolic drive translates into a fortified neurovascular interface, preserving BBB integrity and shielding neural tissues from the metabolic stress of toxic protein accumulation [28, 40, 41]. Similarly, the protective effect observed in oligodendrocyte precursor cells (OPCs) suggests enhanced regenerative capacity supporting myelin maintenance [42, 43]. Our validation using snRNA-seq data from AD patients confirmed the active, disease-associated transcriptional remodeling of these core effectors—including *MAPT* and *MAPT-AS1* [44, 45]—within the glial and pericyte populations, reinforcing the concept that genetically driven NVU fortification acts as a crucial compensatory barrier [46, 47].

These insights carry substantial implications for clinical intervention and drug development.

First, they underscore that aggressive pain management in OA is not solely a matter of preserving mobility, but a critical neuroprotective strategy to prevent accelerated brain aging and cognitive decline [6, 11, 48]. Given the profound genetic pleiotropy shared across skeletal phenotypes and affective disorders, our findings—alongside recent evidence implicating the endocannabinoid system in bone-depression crosstalk [13]—highlight the necessity of a multidisciplinary approach. Effective management must simultaneously address joint nociception and psychiatric comorbidities to preserve cognitive reserve.

Second, our identification of antagonistic pathways serves as a stringent cautionary tale against the uncritical repurposing of therapeutics across the bone-brain axis. Drugs designed to inhibit anabolic pathways (such as PI3K/AKT inhibitors) for the treatment of OA could theoretically compromise neurovascular integrity and inadvertently accelerate neurodegeneration in susceptible individuals [49, 50].

## Conclusions

By applying a multidimensional and cell-type-specific approach, this study resolves the long-standing "bone-brain paradox." We demonstrate that the widely observed epidemiological comorbidity between hip OA and AD is fundamentally a phenotypic illusion, driven by the acceleration of brain aging secondary to chronic pain and the mediating effects of the pain-depression axis [11, 30]. However, beneath this phenotypic overlap lies a profound, genetically anchored antagonism within the bone-brain axis. Our findings reveal that the genetic liability for hip OA confers a robust neuroprotective effect, coordinated by the *MAPT* locus and PI3K/AKT signaling, which specifically fortifies the neurovascular unit (pericytes and astrocytes) [28, 41].

These results yield vital clinical and translational directives. Clinically, they highlight the urgency of effective hip OA pain management as a viable strategy to mitigate depression and decelerate cognitive aging [6, 34]. Translationally, the discovery of this antagonistic pleiotropy mandates extreme caution when developing therapeutic strategies targeting shared molecular nodes within the bone-brain axis, as indiscriminately suppressing OA-associated anabolic pathways may jeopardize the brain’s neurovascular barrier [38, 51]. Ultimately, we propose an evolutionary trade-off paradigm: the systemic mechanisms that predispose the joints to structural vulnerability simultaneously engender protective neurovascular resilience in the aging brain [3, 28, 52].

## Declarations

### Ethics approval and consent to participate

The UK Biobank study was approved by the North West Multi-centre Research Ethics Committee (REC reference: 11/NW/0382), and all participants provided written informed consent prior to data collection. The genome-wide association study (GWAS) summary statistics utilized in this study were obtained from publicly available datasets (UK Biobank, arcOGEN, and EADB). The snRNA-seq data from the Religious Orders Study and Memory and Aging Project (ROSMAP) were collected under protocols approved by the Institutional Review Board of Rush University Medical Center, and all participants signed an informed consent and an Anatomical Gift Act for brain donation. All original studies were conducted in accordance with the Declaration of Helsinki and relevant ethical guidelines, and informed consent was obtained from all participants or their proxies.

### Consent for publication

Not applicable. This study does not contain any individual person’s identifiable data in any form (including individual details, images, or videos).

### Availability of data and materials

The datasets supporting the conclusions of this article are available in the following repositories:

- The GWAS summary statistics for hip osteoarthritis are available in the GWAS Catalog repository (accession no. GCST007091).
- The GWAS summary statistics for Alzheimer’s disease are available in the GWAS Catalog repository (accession no. GCST90027158).
- The single-cell eQTL summary statistics are available in Zenodo (https://doi.org/10.5281/zenodo.5543734).
- The individual-level phenotypic data supporting the observational analyses are available from the UK Biobank database upon application (accession ID 41376).
- The snRNA-seq data from the ROSMAP cohort are available through the AD Knowledge Portal (https://adknowledgeportal.synapse.org/) via Synapse.

### Authors’ contributions

QX conceived and designed the study, performed the data analysis and drafted the manuscript. PZ and JT assisted with the data acquisition, statistical analysis, and interpretation of the results. HZ rigorously revised the manuscript for important intellectual content and supervised the project. All authors read and approved the final manuscript.

## Acknowledgements

This research was conducted using the UK Biobank Resource (accession ID 41376). We thank the participants and researchers of the UK Biobank, the arcOGEN consortium, and the European Alzheimer & Dementia Biobank (EADB) for making their GWAS summary data publicly available. We also extend our gratitude to Bryois et al. for providing the single-cell eQTL data. Furthermore, we gratefully acknowledge the participants of the ROSMAP cohort and the Rush Alzheimer’s Disease Center for providing the single-nucleus transcriptomic data, as well as the AD Knowledge Portal for facilitating data access.

## Additional files

**Additional file 1:** Supplementary tables. **Table S1.** ICD-10 codes and EHR records used for the definition of Hip OA and incident AD. **Table S2.** Univariate and bivariate MiXeR model fit statistics (AIC and BIC). **Table S3.** Detailed parameters of the MiXeR model architecture. **Table S4.** Independent instrumental variables (IVs) selected for hip Osteoarthritis. **Table S5.** Independent instrumental variables (IVs) selected for Alzheimer’s disease. **Table S6.** Full results of the cell-type-specific Mendelian randomization (csMR) analysis across eight brain cell types. **Table S7.** List of 60 distinct pleiotropic lead SNPs identified by conjunctional false discovery rate (ccFDR) analysis.

